# The geno-biomechanical similarity between functional and idiopathic scoliosis from kinematic, muscle activation, vertebral loading and the pathological gene expression perspectives

**DOI:** 10.1101/2025.11.08.25339358

**Authors:** Baichun Wei, Zihang Xu, Jianfei Zhu, Kai Zhu, Haijiao Wang, Chao Gan, Jinnan Duan, Zuowei Jiang, Xun Wang, Jin Cui, Jin Tian, Yufeng Wang, Yuli Fang, Lijun An, Xiaolei Sun, Chunzhi Yi

## Abstract

Adolescent functional scoliosis arises from complex interactions among inappropriate habits, incorrect force field induced by asymmetric muscle activation and potentially abnormal gene expression during development. Growing evidence suggests that untreated functional scoliosis may progress to idiopathic scoliosis, while debatable in the latest SOSORT guidelines for American adolescents. To characterize the biomechanical mechanisms underlying this potential progression, we integrated multi-modal data, including kinematics, muscle activation, vertebral loading and gene expression to comprehensively understand how subjects with functional scoliosis presents similarity with patients with idiopathic scoliosis compared with healthy controls. The results from 33 subjects with functional scoliosis, 27 subjects with idiopathic scoliosis and 18 healthy controls show that subsets of subjects with functional scoliosis present a closer kinetics, muscle activation pattern to those with idiopathic scoliosis compared with healthy controls, according to the comprehensively assessed features from inertial measure unit (IMU) and surface myoelectric (sEMG) signals. Subject-specific musculoskeletal modelling also shows a closer pattern in bone loading between functional and idiopathic scoliosis compared with healthy controls. The genetic expression analysis further indicates a significant correlation between the phenotype of functional scoliosis with the highest pathological information and the pathological genetic mutation for idiopathic scoliosis. Our results provide insights into the complex interaction across kinetic, biomechanical and genetic aspects of functional and idiopathic scoliosis, suggesting that a subset of functional scoliosis may represent early manifestations of idiopathic scoliosis. These insights may inform future updates to guidelines for the management of functional scoliosis.

## Introduction

Adolescent idiopathic scoliosis (AIS), definined as a lateral spinal curvature with a Cobb angle ≥10° in individuals aged 10-18 years**Error! Reference source not found.**, is the most common form of scoliosis **Error! Reference source not found.Error! Reference source not found.**, with a higher prevalence in females than males **Error! Reference source not found.**. AIS is a three-dimensional deformation of spine slowly progressed with growth, resulting in bending or twisting of vertebra with unknown reasons. AIS has been thought as a syndrome with multi-factorial origins and its etiopathogenesis (the answer of “Which force drives scoliotic deformation of spine” **Error! Reference source not found.**) has not been elucidated **Error! Reference source not found.**. As a result, despite screening AIS patients has been widely adopted in early detection and can help to slow disease progression by introducing early intervention **Error! Reference source not found.**, predicting and preventing the onset of AIS based on some early signs are still largely under-explored.

Functional scoliosis, a non-structural, mild, steady curvature without vertebra rotation, is usually treated as a separate type of scoliosis in addition to idiopathic scoliosis **Error! Reference source not found.**. According to the recommendation of SOSORT guidelines, clinical procedures of functional scoliosis is suggested with observation and lowest-level treatment **Error! Reference source not found.**. Indeed, there lacks evidence on how functional scoliosis may progress into idiopathic scoliosis, despite recent studies showing the susceptibility to AIS of adolescents even with incorrect postures **Error! Reference source not found.**-**Error! Reference source not found.**. Theoretically, asymmetric muscle activation **Error! Reference source not found.**-**Error! Reference source not found.** induced by abnormal curvature of spine and reduction of muscle mass **Error! Reference source not found.Error! Reference source not found.** that usually associates with scoliosis lead to a pathological force environment of vertebra and intervertebral discs. Such force environment may induce abonormal gene expression and imbalanced spinal growth compared with ligament and other soft tissues **Error! Reference source not found.**-**Error! Reference source not found.**, thus may escalate into vertebra rotation [5]. Here, we fill in the knowledge gap by providing geno-biomechanical evidence on how functional scoliosis patients show stronger similarity with AIS patients compared with healthy subjects.

Specifically, we integrated multi-modal wearable sensors, individualized musculoskeletal modelling and gene expression data to compare the kinematics, muscle activation, spinal loading and RNA expression levels in blood and paraspinal muscles across healthy, functional and idiopathic scoliosis adolescents. We showed that functional scoliosis presented unique kinematic pattern during walking, a closer muscle activation and vertebral loading pattern with idiopathic scoliosis during walking and Adam’s forward bending test. All the phenotypes of functional scoliosis presented stronger associations with gene expression of idiopathic scoliosis, compared with the phenotypes of healthy controls. These suggest that functional scoliosis may be an early sign of idiopathic scoliosis and should be paid more focus during clinical treatment.

## Results

### Overview

We performed the multi-modal data analysis in following steps. We recruited 33 subjects with functional scoliosis, 27 subjects with idiopathic scoliosis and 18 healthy controls (see “Methods-Participants”) to perform two tasks, shuttle walking and Adam’s forward bending test (see “Methods-Motion tasks”), with wearable IMU, EMG and foot pressure sensors (see “Methods-Sensor placement” and Fig. S1 and S2 for sensor placement details). The subjects also walked on ground force plates with reflective markers under optical motion capture system to develop individualized musculoskeletal model (see “Methods-Individualized musculoskeletal model”). We extracted EMG features in both individual muscle level and muscle synergy level from data collected during shuttle walking and forward bending, kinematic features from data collected during shuttle walking and vertebral loading from musculoskeletal models (Fig. 1a). We analyzed similarities between each pair of the three groups of subjects through multiple methods (Fig. 1b) and then compared the similarity between functional scoliosis and healthy controls with that between functional and idiopathic scoliosis. We also associated EMG features with kinematic features and vertebral loading to investigate whether the EMG feature screened by decoder-based analysis can explain the variance of kinematics and spinal loading. We calculated the correlation between the three kinds of phenotypes and gene expression in blood and paraspinal muscles of idiopathic scoliosis patients. This is to show the differential association between the phenotype of functional scoliosis and idiopathic scoliosis compared with healthy subjects.

**Fig 1.**
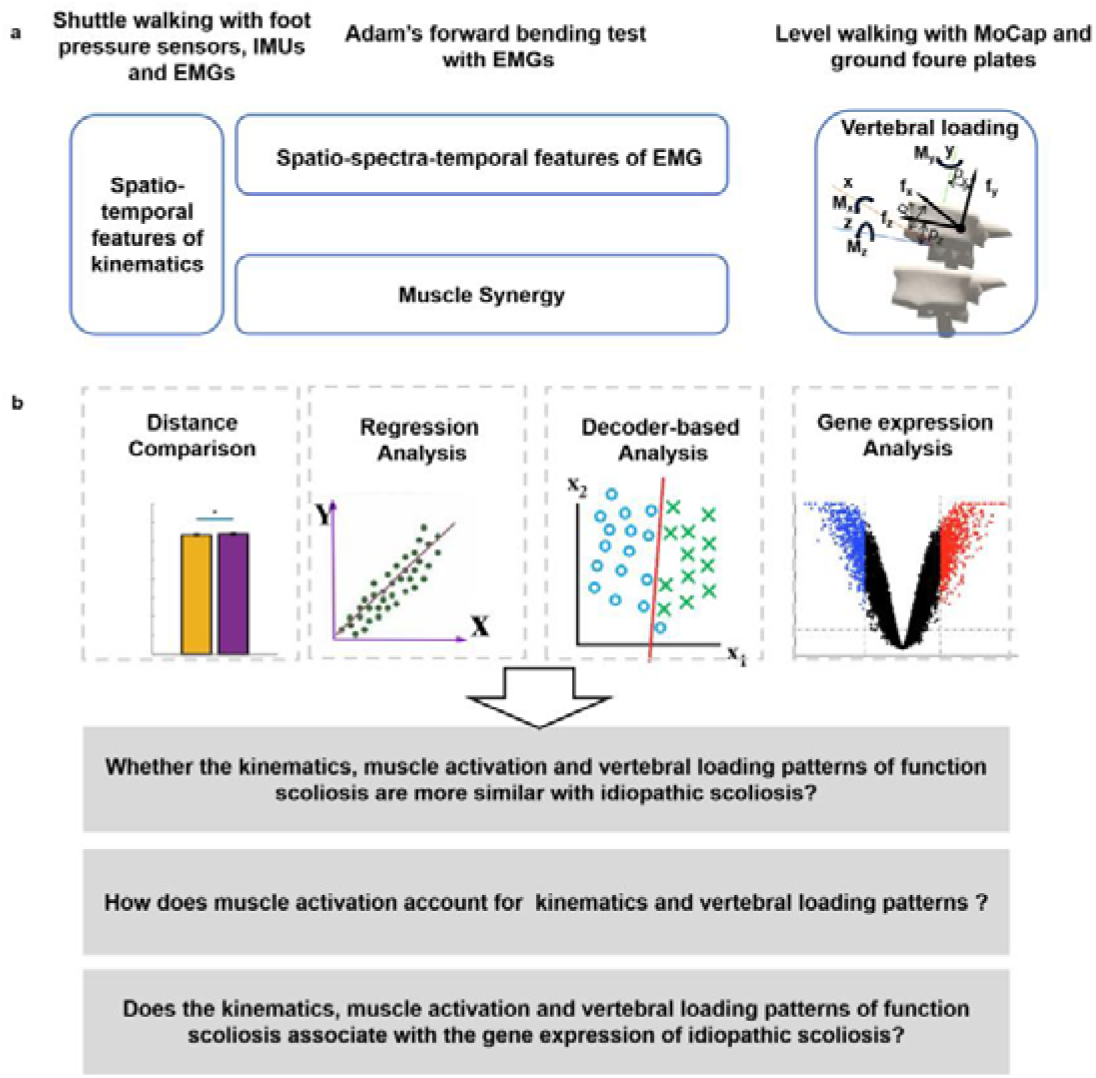
Overview of the data analysis pipeline. a We collected the IMU, EMG and foot pressure signals during shuttle walking and EMG signals during Adam’s forward bending test. We collected optical motion capture and ground force plate data during walking (See “Methods-motion tasks and sensor placement”). We extracted spatio-temporal features from IMU signals during shuttle walking, spatio-spectra-temporal features of each muscle and muscle synergy features from EMG signals during shuttle walking and forward bending, as well as vertebral loading enabled by individualized musculoskeletal modelling. b We then compared the distance of each modal among the three groups of subjects, regressed among groups to show co-variance, subgrouped functional scoliosis patients based on linear decoder and analyzed the associations of the phenotypes with gene expression of idiopathic scoliosis patients to answer the three questions shown in the figure.

### Muscle activation of functional scoliosis subjects present closer pattern to that of idiopathic scoliosis patients

We first tested whether the muscle activation patterns during walking and Adam’s test of functional scoliosis were closer to idiopathic scoliosis, compared with healthy controls. If so, the similarly asymmetric force field constructed by the muscles surrounding spine would on one hand affect the asymmetry of vertebral loading, induce a further asymmetry of the spinal curvature through stimulating misexpression of genes as well as inelastic deformation, thus may deteriorate the pathology. One the other hand, the similar muscle activation pattern suggests potentially similar pattern of gene mutation or abnormal expression on DNA methylation as well as small non-coding RNA, which would drive the 3D deformation of spine during growth. To evaluate this, we extensively calculated the EMG features from the 6 muscles surrounding spine and 8 lower-limb muscles for walking and the same 6 muscles surrounding spine and for Adam’s forward bending test (see “Methods-Sensor placement”). The features spanned temporal, spectral and spatial domains (See “Methods-EMG features and Kinematic features” and SI 1 and 2).

We constructed the distribution of the maximum mean discrepancy between each pair of the three groups (functional scoliosis, healthy controls and idiopathic scoliosis) using bootstrap methods (Fig. 2a, see “Methods-Statistics”) and then compared across the pairs of groups (Fig. 2b and c). We found that the distance for walking between functional scoliosis and healthy controls is similar to that between functional and idiopathic scoliosis, and smaller than that between healthy controls and idiopathic scoliosis. The distance for Adam’s forward bending test between functional scoliosis and healthy controls is larger than that between functional and idiopathic scoliosis (Fig. 2c). This indicates a heterogeneous pattern between postures and a closer muscle activation pattern, in temporal, spatial and spectral domains, between functional and idiopathic scoliosis. Next, we regressed the EMG features of functional scoliosis against those of healthy controls and idiopathic scoliosis (see “Methods-Statistics”), respectively. We found that the covariance, denoted by the regression coefficient of the linear regression, is larger between functional and idiopathic scoliosis (Fig. 2d). We then used C-SVM, a linear decoder, to classify the three groups of subjects using their EMG features (Fig. 2e, see “Methods-Machine learning-based analysis”). We subgrouped the subjects with functional scoliosis that were classified as different groups (denoted by different predicted labels in Fig. 2f and g) and regressed their EMG features against those of healthy subjects and idiopathic scoliosis patients, respectively. By comparing the regression coefficients across predicted labels, we found subjects with functional scoliosis present heterogeneous similarity (Fig. 2f and g). Some subjects within this group showed significantly high similarity with both healthy subjects and idiopathic scoliosis patients (the subgroup with the label of “Healthy”). The subgroup with the label of “Functional” showed a higher similarity with idiopathic scoliosis patients. This indicates a heterogeneous distribution of the muscle activation within these functional scoliosis subjects, suggesting a potential subgrouping requirement. By using both C-SVM and logistic regression, we screened the distinguishable EMG features across the three groups of all the subjects as the alpha band coherence between left and right paraspinal muscles beside thoracic vertebra for Adam’s forward bending test (see “Methods-Machine learning-based analysis”). Taken together, the muscle activation pattern assessed by temporal, spectral and spatial EMG features across different muscles and different postures showed that the subjects with functional scoliosis present to be similar to the patients with idiopathic scoliosis, compared with healthy controls.

**Fig. 2.**
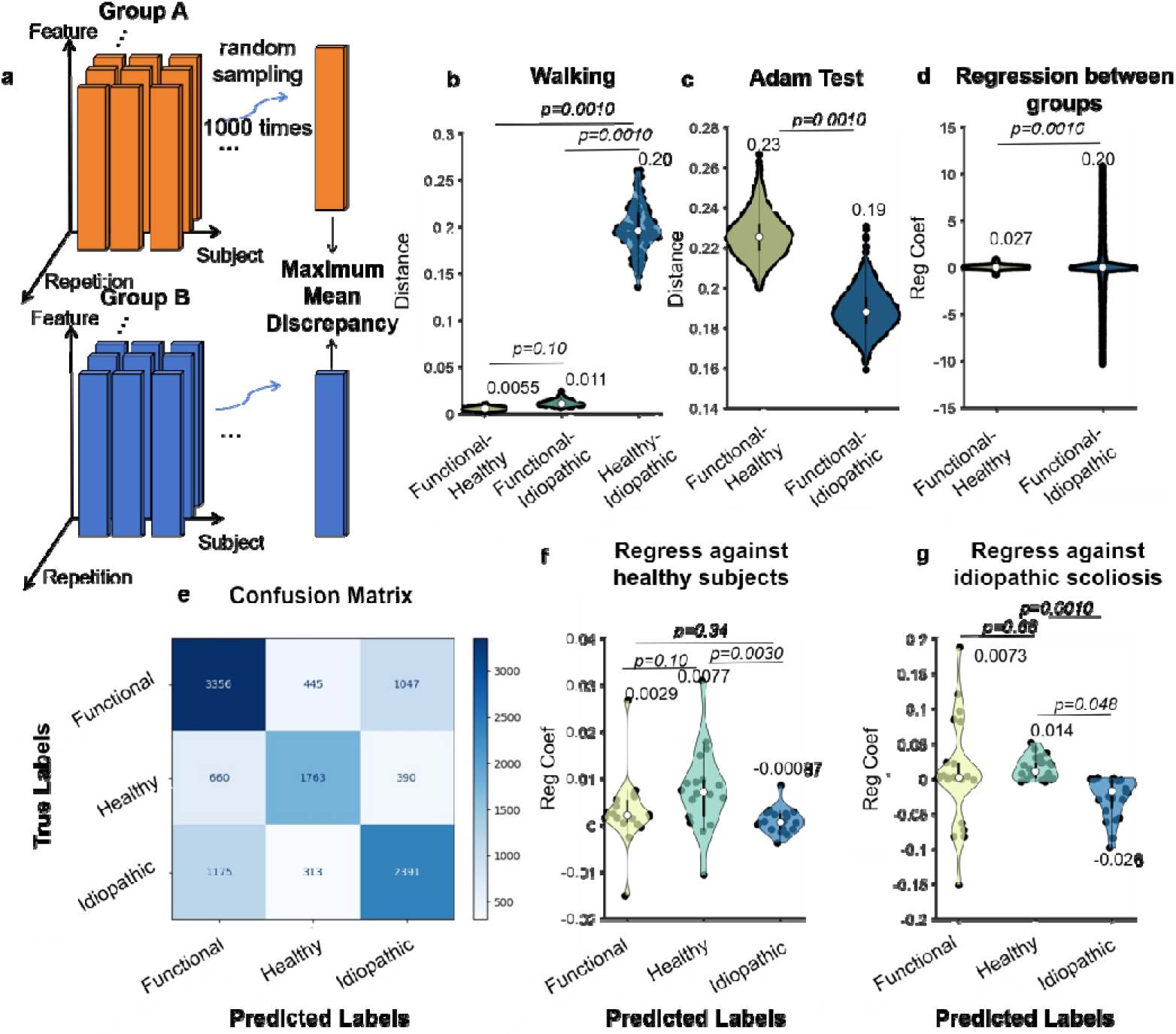
Muscle activation pattern analysis. We randomly sampled features from separate groups and calculate the maximum mean discrepancy (MMD) for 1000 times (a). We compared MMD between each pair of the three groups for walking (b) and Adam’s test (c). We performed linear regression between functional scoliosis and healthy controls as well as between functional and idiopathic scoliosis and compared the regression coefficients (d). We used c-SVM to classify the three groups using EMG features (e) and compared the regression coefficients of subgroups within functional scoliosis (Predicted Labels) with healthy subjects (f) and idiopathic scoliosis patients (g). All the p values were corrected for multiple comparisons using FDR method.

### Muscle synergy pattern of walking showed a greater similarity between functional and idiopathic scoliosis

We turned to evaluating how muscles coordinate with each other in the three groups. Muscle synergy typically assessed by non-negative matrix factorization has been thought to reflect the compact motor command from central nervous system and the synergistic nature of some low-level discrete elements within motor system. We then assessed the similarity of muscle synergy across groups and the the sparseness. We found the muscle synergy of the subjects with functional scoliosis during walking was more similar to that of the patients with idiopathic scoliosis, compared with healthy controls (Fig. 3a, see “Methods-EMG features”, also SI 1). The sparseness of muscle synergy for functional scoliosis was similar to idiopathic scoliosis and lower than that of healthy controls (Fig. 3b, see “Methods-EMG features”, also SI 1). This suggested the spinal curvature induced by both functional and idiopathic scoliosis induced muscle compensation so that muscles showed an enlarged co-activation involving redundant muscles. For Adam’s forward bending test, no significant difference was found in similarity and sparseness (Fig. 3c and d).

**Fig. 3.**
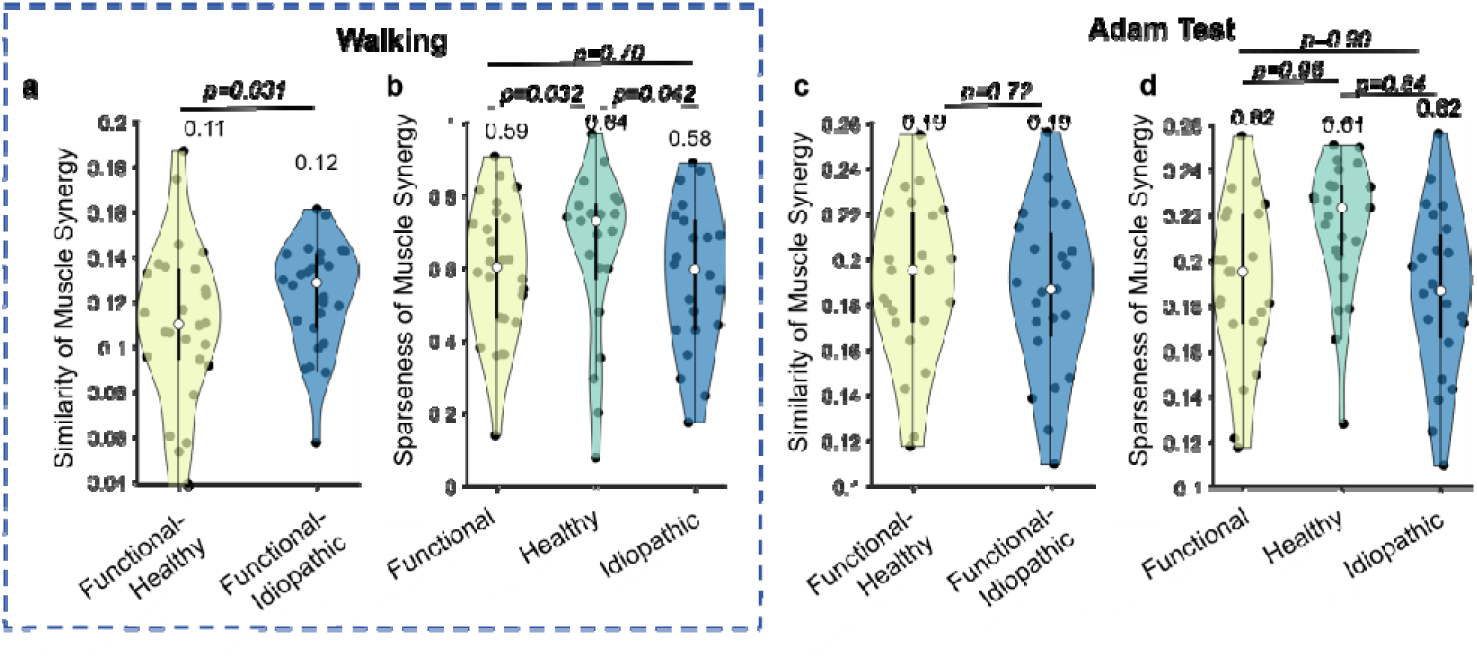
Analysis on muscle synergy across the three groups for shuttle walking and Adam’s forward bending test. Similarity of muscle synergy for walking (a) and Adam’s forward bending test (c), as well as the sparseness of muscle synergy for walking (b) and Adam’s forward bending test (d). All the p values were corrected for multiple comparisons using FDR method.

### Functional and idiopathic scoliosis disturbed the association between muscle activation patterns and kinematic features

Activated muscles drive motions and form kinematic output. After evaluating the muscle activation patterns across groups, we turned to evaluating whether kinematics showed consequent variations, despite the many-to-many mapping between muscle activation and kinematics. We tested whether the kinematic features of functional scoliosis subjects were more similar to those of idiopathic scoliosis patients and how they associated with muscle activation patterns. We assessed kinematics during shuttle walking through extensive features from IMUs (see “Methods-Kinematic features”). We found that the distance of between functional scoliosis and healthy controls is similar to that between functional and idiopathic scoliosis, which is larger than the distance between healthy controls and idiopathic scoliosis (Fig. 4a). This suggests a unique kinematic pattern for functional scoliosis during walking. Moreover, the regression coefficients showed that the kinematic features of functional scoliosis had a covariance with healthy controls greater than that with idiopathic scoliosis (Fig. 4b). These results suggested that despite the muscle activation and synergy patterns of functional scoliosis closer to those of idiopathic scoliosis, the many-to-many mapping between synergistic muscle activation and kinematics led to a greater similarity between functional scoliosis and healthy controls. We then trained logistic regression model to classify the three groups using kinematic features. We showed that the subjects with functional scoliosis classified as healthy controls (“Predicted Labels” in Fig. 4d and e, see “Methods-Machine learning-based analysis”) showed greater similarity with both healthy controls and idiopathic scoliosis patients than the rest two subgroups. Moreover, the similarity between the subjects with functional scoliosis classified as idiopathic scoliosis (denoted by idiopathic scoliosis subgroup) and healthy controls as well as the similarity between idiopathic scoliosis subgroup and idiopathic scoliosis was greater than the similarity for functional scoliosis subgroup (Fig. 4d and e). These results again suggested the potential confusion caused by the many-to-many mapping between muscle activation and kinematics. To test the association between muscle activation patterns and kinematic features, we regressed the first principle components of EMG features against the first principle components of kinematic features, where the first principle components were obtained by performing principle component analysis (PCA). Only healthy controls showed a significant association between EMG and kinematic features after controlling gender and curve pattern (Fig. 4f), suggesting both functional and idiopathic scoliosis may disturb the mapping between activation of multiple muscles and kinematics. Then, we identified screened features by finding the intersect between the top features selected by the weight of logistic regressor and the top features selected by the accuracy degradation in C-SVM-based analysis after permuting each feature for 1000 times (see “Methods-Machine learning-based analysis”). The screened features were single support phases of right and left feet, pitch of the IMU mounted on left thigh and yaw of the the IMU mounted on left shank for kinematic features (IMU placement refer to Fig. S1 and S2). We found the screened features showed a greater association with EMG features, compared with other kinematic features (Fig. 4g, see “Methods-Statistics”). Together, we found the association between muscle activation patterns and kinematic features was disturbed by both functional and idiopathic scoliosis, despite the many-to-many mapping between them led to a similar distribution of kinematic features between functional scoliosis and healthy controls.

**Fig. 4.**
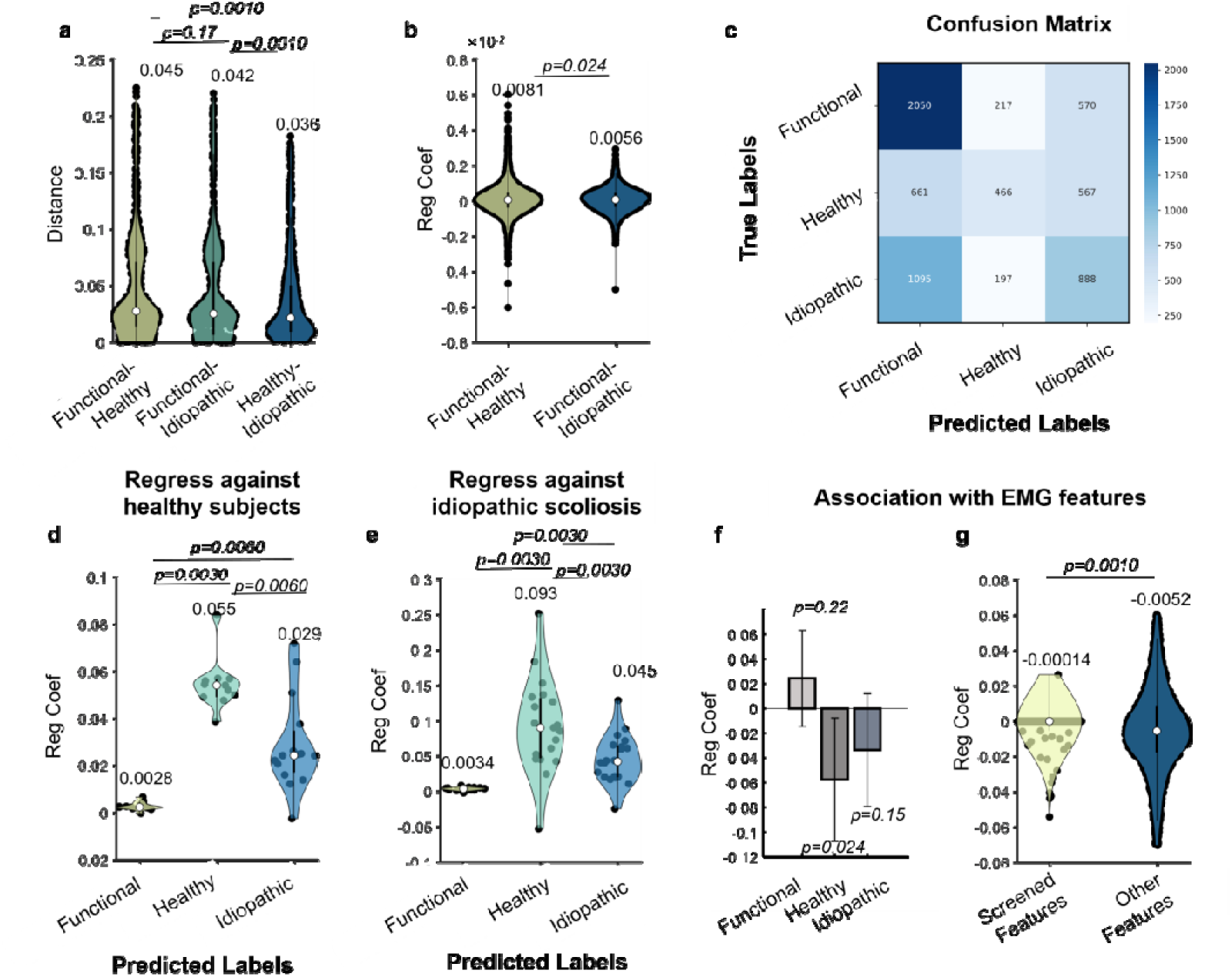
Analysis on kinematic features across the three groups for walking. We calculated the MMD between each pair of the groups and constructed the distribution by bootstrapping (a). We regressed the kinematic features of functional scoliosis subjects against those of healthy controls and idiopathic scoliosis patients (b). We classified the three groups using kinematic features through logistic regression model (c) and subgrouped the subjects with functional scoliosis and compared the similarity between each subgroup with healthy controls (d) and idiopathic scoliosis patients (e). We regressed the first principal components of EMG features against that of kinematic features (the first principal components of kinematic features ∼ gender + age + curve pattern + the first principal components of EMG features) (f). We identified the screened features from kinematic features and then compared their association with EMG features and the association of the rest kinematic features with EMG features (the first principal components of kinematic features ∼ gender + age + curve pattern + the first principal components of EMG features) (g). All the p values except for those in f were from permutation test corrected for multiple comparisons using FDR method.

### The vertebral loading associated with muscle activation and presented a greater similarity between functional and idiopathic scoliosis

The long-lasting asymmetric force field constructed by abnormal muscle activation and the already existed spinal curvature may cause stress concentration and may lead to abnormal gene expression induced by the mechanical environment. In the following, we asked whether the vertebral loading within the functional scoliosis subjects showed similar pattern with the idiopathic scoliosis patients. To do this, we assessed the vertebral loading through individualized musculoskeletal model built from X-ray images of spine as well as the data from motion capture system and ground force plates (Fig. 5a, see “Methods-Individualized musculoskeletal model”and Fig. S3). We performed the same procedure as reported in kinematic features session to identify the screened features from EMG features (see “Methods-Machine learning-based analysis”). The screened EMG features were RMS ratio between left and right quadriceps femoris, alpha band power of left gluteus medius, beta band power of right musculus gastrocnemius and RMS of right musculus gastrocnemius (corresponding to the leftmost four bars in X axis of Fig. 5c) for shuttle walking. Then, we showed that the screened EMG features for all the three groups show significant association with the first principal component of the vertebral loading after controlling gender and curve pattern (Fig. 5c, see “Methods-Statistics”). We then showed that the distance between functional and idiopathic scoliosis is smaller than that between functional scoliosis and healthy controls (Fig. 5b), indicating the greater similarity on the pattern of vertebral loading between functional and idiopathic scoliosis.

**Fig. 5.**
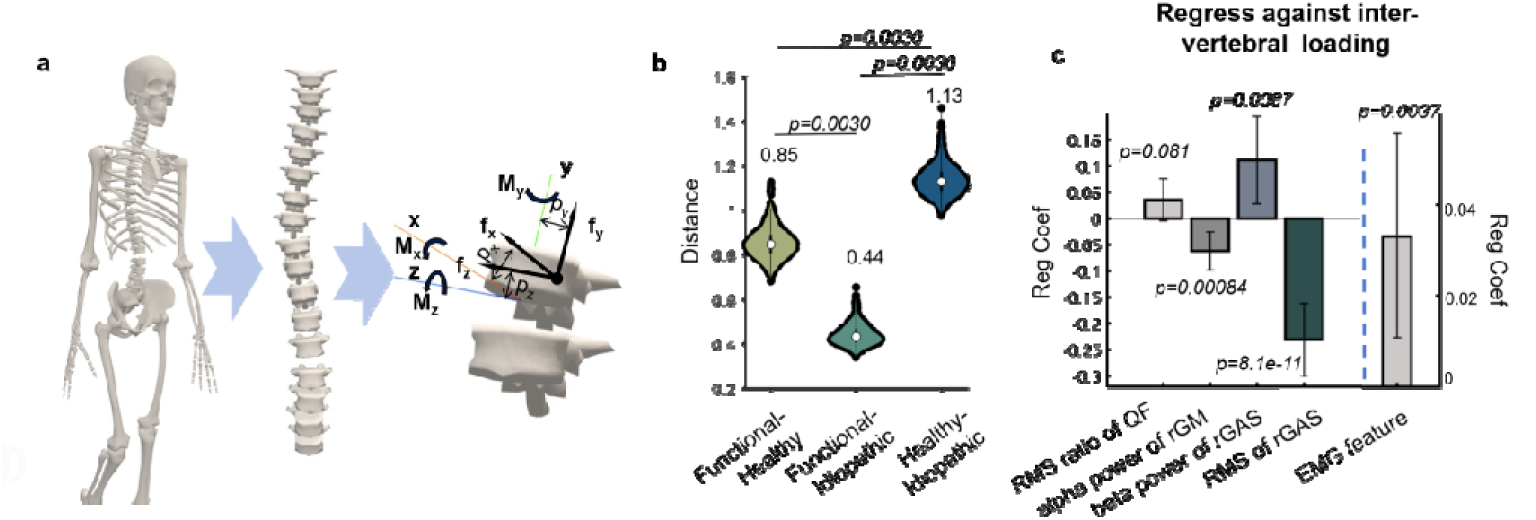
Analysis on vertebral loading during walking enabled by individualized musculoskeletal modelling. We built individualized musculoskeletal models according to the X-ray images and data from motion capture and ground force plates of each subject then evaluated the loading of each vertebra (a). We constructed the MMD distribution and compared across the pairs of the three groups (b). The p values in (b) were corrected for multiple comparisons using FDR method. We regressed the screened features of EMG features (the left four bars in c) and the first principal component of EMG features (the rightmost bar in c) against the first principal component of vertebral loading (the first principal component of vertebral loading ∼ the first principal component of EMG features or the screened EMG features + age +gender + curve pattern).

### The abnormal patterns of kinematics, muscle activation and vertebral loading for functional scoliosis were associated with the expression of pathogenic gene of idiopathic scoliosis

Previous studies have indicated the abnormal expression of genes may modulate the abnormal growth of vertebra and intervertebral disc during growth for idiopathic scoliosis patients and thus may affect kinematics and muscle activation patterns. We have assessed the similarities of vertebral loadings among functional scoliosis, healthy controls and idiopathic scoliosis, as well as the association between vertebral loading and muscle activation patterns. In the following, we turned to assessing how the gene expressions of paraspinal muscles and blood from idiopathic scoliosis patients associate with the kinematics, muscle activation and vertebral loading patterns of functional scoliosis patients. We asked the question of whether the correlation between the pathological generic expression and the phenotype, including kinematics, muscle activation and vertebral loading, is differential across healthy control, functional and idiopathic scoliosis. We correlated kinematic, EMG features and vertebral loading patterns (Fig. 6a, b and c, see “Methods-Gene expression analysis”) of healthy controls, functional and idiopathic scoliosis patients recruited in our study with the mRNA within paraspinal muscles of idiopathic scoliosis patients**Error! Reference source not found.**. We found that the correlations for both kinematics features and vertebral loading patterns of functional scoliosis patients were different from those of healthy controls (Fig. 6a, p = 0.044; Fig. 6 c, p=0.0010), while similar to those of idiopathic scoliosis patients (Fig. 6a, p=0.63; Fig. 6c, p=0.83).

**Fig. 6.**
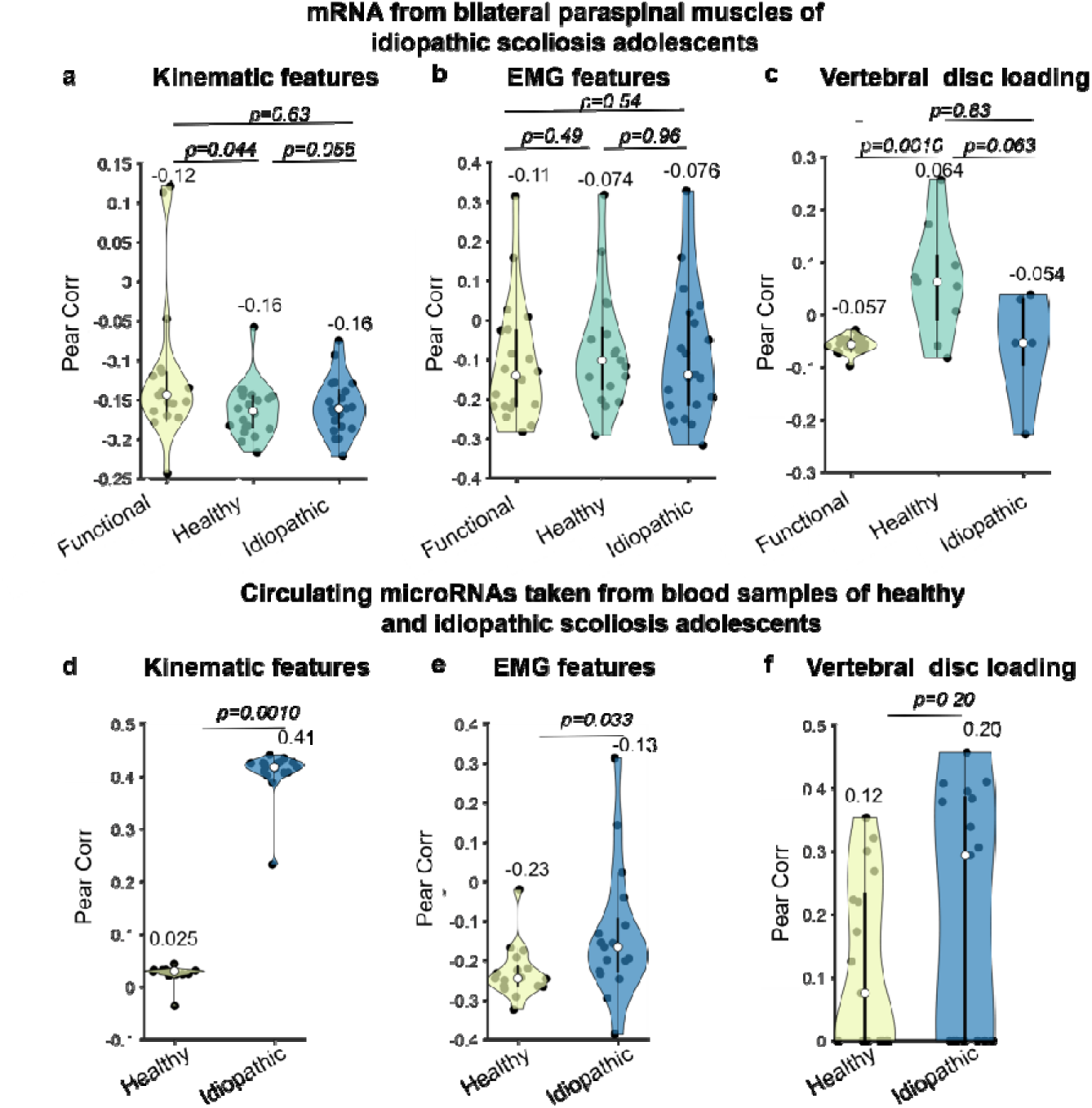
Similarity comparison on gene expression among healthy control, functional and idiopathic scoliosis groups. The first row denotes the Pearson’s correlation coefficients between the first principal components of each phenotype (a for kinematic features, b for EMG features and c for intervertebral disc loading) from all the three groups of people we recruited and the first principle component of mRNA of bilateral paraspinal muscles taken from idiopathic scoliosis patients during surgery. The X axes denote each group of subjects recruited in our study. The second row denotes the Pearson’s correlation coefficients between the first principle components of each phenotype (d for kinematic features, e for EMG features and f for intervertebral disc loading) from functional scoliosis patients and the first principal components of circulating microRNA taken from blood samples from healthy and idiopathic scoliosis subjects. The X axes denote healthy and idiopathic scoliosis patients for taking blood samples.

To complement the findings above, we further correlated the kinematic, EMG features and intervertebral disc loading patterns (Fig. 6d, e and f, see “Methods-Gene expression analysis”) of functional scoliosis patients with the circulating microRNA taken from blood samples of idiopathic scoliosis patients and healthy subjects**Error! Reference source not found.**. We found that the correlations for the kinematic and EMG features of functional scoliosis patients recruited in our study presented to be larger with circulation microRNA of idiopathic scoliosis patients than with circulation microRNA of healthy subjects (Fig. 6 d and e). Taken together, the findings indicated that the phenotype including kinematics, muscle activation and spinal loading for functional scoliosis patients showed closer correlation with the gene expression of idiopathic scoliosis, compared with healthy subjects. The potential link between the phenotype of functional scoliosis patients and abnormal gene expression for idiopathic scoliosis further suggested, combining with the findings of all the sessions, functional scoliosis might be an early sign of idiopathic scoliosis.

## Discussion

Functional and idiopathic scoliosis has been largely considered as separate types of scoliosis. Our study shows geno-biomechanical evidence that the functional scoliosis patients present closer muscle activation, spinal loading and generic associations with idiopathic scoliosis patients, compared with healthy controls, despite functional scoliosis patients show unique kinematic patterns. This essential cross-scale similarity indicates the musculoskeletal pattern during walking and bending forward (the most common motion and the distinguishable motion for diagnosis) for functional scoliosis underlying kinematics show an essential shift toward idiopathic scoliosis, suggesting the potential transformation from functional scoliosis into idiopathic scoliosis. The findings provide novel insight into the clinical practice of treating functional scoliosis as a potential early sign of AIS.

We add the conceptual advance to the literature on the musculoskeletal association between functional and idiopathic scoliosis compared with healthy subjects. Our study is grounded in the experience-based observations showing the susceptibility of even bad or incorrect postures towards idiopathic scoliosis[11]**Error! Reference source not found.**[25]. Also, a recent study has shown that the leg length inequality associated with functional scoliosis corresponded to vertebra rotation**Error! Reference source not found.**, which challenges the traditional idea that functional scoliosis only involves two-dimensional deformity of spine. To the best of our knowledge, there is no study showing the biomechanical evidence on the association between functional and idiopathic scoliosis. Our findings further indicate the biomechanical similarity underlying kinematics between functional and scoliosis from the perspectives of muscle activation, coordination patterns, spinal loading patterns and their association with gene expression of idiopathic scoliosis patients, which provide further demonstrations on the experience-based observations. Also, our findings elucidate the possibility of the deterioration from functional into idiopathic scoliosis.

Such findings meet the long-lasting assumption of the differential growth between vertebra and ligament**Error! Reference source not found.Error! Reference source not found.** induced by the imbalanced force field of muscles and tendons, as well as the intuition that any deformity is associated with mechanical force[5]. Following this assumption, functional scoliosis induces imbalanced muscle force performed on spine and thus uneven distribution of spinal compression. According to Hueter-Volkmann law**Error! Reference source not found.**-**Error! Reference source not found.**, the consequently insufficient spinal compression at some area of vertebra, which may be further deteriorated by relatively low muscle mass**Error! Reference source not found.Error! Reference source not found.**, would lead to decreased inhibition of vertebra and intervertebra disc growth, thus resulting in abnormally overgrowth of vertebra as observed in AIS patients**Error! Reference source not found.**-**Error! Reference source not found.**. Moreover, the ligament growth that usually relies on dynamic loading**Error! Reference source not found.** cannot be facilitated by the static and quasi-static loading**Error! Reference source not found.**, thus may not match the growth of vertebra during puberty and induce twisting and three-dimensional deformity of spine. As demonstrated by a physical model**Error! Reference source not found.**, subtle differential growth would induce a scoliotic deformation.

We add the knowledge advance to the literature on showing the kinematics, muscle activation and muscle synergy patterns of functional scoliosis in the context of comparing them with those of idiopathic scoliosis and healthy subjects. Our findings also extend previous findings on idiopathic scoliosis and other structured scoliosis to functional scoliosis. On kinematics, previous studies have indicated the key kinematic features showing significant association with spinal curvature of scoliosis**Error! Reference source not found.** and significant changes comparing with healthy subjects**Error! Reference source not found.**. Our study further adds the key kinematic features that are distinguishable across functional, idiopathic scoliosis and healthy subjects, which details the previous findings. Our findings indicate the motion of left thigh, shank and single support phases for both feet as the key gait kinematic features from extensive spatio-temporal gait features through joint utilization of two linear machine learning-based decoders. This generally meets previous findings in distinguishing scoliosis and healthy subjects, but provides further details. On muscle activation, by extensively comparing the spectra-spatio-temporal EMG features across the three groups in our study, our findings indicate the alpha band coherence between right and left paraspinal muscles beside thoracic vertebra for forward bending and RMS ratio between left and right quadriceps femoris, alpha band power of left gluteus medius, beta band power of right musculus gastrocnemius and RMS of right musculus gastrocnemiu for walking as the distinguishable features across functional, idiopathic and healthy subjects. The findings generally meet the findings from previous studies in scoliosis or AIS**Error! Reference source not found.**-**Error! Reference source not found.** and provide details in the muscle activation difference specific to the three groups of subjects recruited in our study. On muscle synergy, we introduce the muscle synergy similarity and sparseness into this field, which has been used and demonstrated its efficiency in sports**Error! Reference source not found.**. All of the screened features may assist to develop digital markers for early diagnosis, early warning or disease progression prediction for functional and idiopathic scoliosis.

We also add the knowledge advance to the literature that the kinematic, muscle activation and vertebra loading patterns of functional and idiopathic scoliosis are similarly associated with gene expression of idiopathic scoliosis. And these patterns of functional scoliosis are more associated with the gene expression of idiopathic scoliosis compared with the gene expression of healthy subjects. Previous studies have shown differentially expressed gene across AIS, healthy subjects and other scoliosis patients**Error! Reference source not found.**[23] as well as their associations with clinical measures (e.g. Cobb angle, curve type**Error! Reference source not found.** age of onset**Error! Reference source not found.**), imaging metrics (e.g. fat infiltration, muscle atrophy**Error! Reference source not found.**), and molecular signatures (e.g. mechanotransduction in osteoblasts**Error! Reference source not found.**, plasma proteomics**Error! Reference source not found.**). Our findings link the kinematics, muscle activation, spinal loading and gene expression to show the potential geno-biomechanical pathway underlying the observations across functional scoliosis and idiopathic scoliosis. Our findings provide mechanistic insights into the potential link and even transformation between these two types of scoliosis, which may guide the design of future work sin animal models or molecular studies.

Our findings may provide some insights into the clinical practice. First, physiotherapeutic scoliosis-specific exercise (PSSE) may benefit the functional scoliosis patients and may impede their disease progression. Despite the SOSORT guideline suggested an observation with periodic follow-up for the idiopathic scoliosis patients with Cobb angle lower than 20° during growth as the main clinical treatment, previous studies have indicated the benefits of PSSE on treating such patients**Error! Reference source not found.**. Our findings provide geno-biomechanical details and possible mechanisms underlying such evidence-based observations. Second, the machine learning-based subgrouping of functional scoliosis patients shows differential similarities with healthy and idiopathic scoliosis patients, indicating the possibility of subtypes within functional scoliosis. Further studies on large cohort may help to determine the biological and digital biomarkers of such functional scoliosis, which may consequently lead to more subtle treatment strategies.

We add the methodological advance by providing a framework utilizing multi-modal signals from wearable sensors, medical image-enabled individualized musculoskeletal model and gene expression analysis, which can be generalizable in multiple fields. We collected signals from wearable signals through two motion tasks, one for the most common daily motion and the other for the mostly used motion for diagnosis. We performed distribution distance, linear regression and decoder-based analysis to evaluate the similarity across groups separately for kinematics, muscle activation and spinal loading. The individualized musculoskeletal models were developed according to the recommendation of recent study**Error! Reference source not found.** and further calibrated by the motion capture data collected during walking (i.e. the specific motion task for analyzing the spinal load). In this way, the biological plausibility of the models can be maximally guaranteed. We then regressed the EMG features against kinematic features and vertebra loading features, respectively, to investigate how the muscle activation can explain the variance of kinematics and spinal loading. Finally, we recruited gene expression of idiopathic scoliosis and associated them with the biomechanical patterns. We anticipate this cross-scale framework can be applied in analyzing and predicting sports injury, efficiency of orthopedic treatment and assistive robots.

Our study has several limitations. First, the limited amount of participants may constrain the finding of more distinguishable features of kinematics and muscle activation. Second, the gene expression data were open-sourced and cannot correspond to our data person to person, which limits in-depth analysis. Third, more biologically detailed musculoskeletal model**Error! Reference source not found.** can be used to provide more detailed and physiological explanation of our results. Finally, a longitude cohort needs to be built to further track the biomechanical development of functional scoliosis patients.

## Methods

### Participants

The study was approved by the fifth hospital of Harbin Ethics Committee (KY2023-01). We recruited participants at their first presentation at our clinic between June 2023 and September 2023. For the healthy controls, we utilized Adam’s forward bending test trunk rotation angle measured with a scoliometer, and a radiation-free spine ultrasound imaging system (Scolioscan® SCN201, Hong Kong) to ensure there were no scoliotic deformities. For functional scoliosis patients, the inclusion criteria were as follows: (1) Cobb angle between 10^。^ and 15^。^; (2) ATR<5^。^. The exclusion criteria were as follows: (1) the Cobb angle and ATR fall out of the inclusion criteria; (2) other types of scoliosis (e.g. congenital scoliosis, neuromuscular scoliosis, etc; (3) leg length inequality. For idiopathic scoliosis, the inclusion criteria were as follows: (1) Cobb angle between 20^。^and 45^。^; (2) ATR≥5^。^; (3) removing orthosis over 24 hours before collecting data. The exclusion criteria were as follows: (1) do not fit the inclusion criteria; (2) other types of scoliosis (e.g. congenital scoliosis, neuromuscular scoliosis, etc). Participants of all the abovementioned three groups should be 10-18 years old. After screening the eligibility of 200 adolescents, we adopted 33 subjects with functional scoliosis, 27 subjects with idiopathic scoliosis and 18 healthy controls (demoscracy and curvature information refer to Tab. S1-S3).

### Motion tasks

The participants performed two motion tasks, i.e. level waking and Adam’s tests. For level walking, the participants walked 10 m with self-determined speed wearing IMU, EMG and foot pressure sensors repeatedly for 10 times. When analyzing the level walking data, we removed the data when the participants turned around by video and foot pressure signals. For Adam’s forward bending test we asked the participants to perform Adam’s forward bending test for 5 seconds, where the beginning and the end of the test were manually labeled by pressing the foot pressure sensor. When analyzing the data of Adam’s forward bending test we removed the data during the first and last 1 second.

### Sensor Placement

We placed sensors in different manners across motion tasks. For level walking, we collected EMG signals (Trigno WirelessSystem; DELSYS, Boston, MA, USA, sampling rate at 1111.11Hz) from three pairs of paraspinal muscles beside T3, T8 and L1. We also collected EMG signals from quadriceps femoris, tibialis anterior, gastrocnemius and gluteus medius muscles on both sides. We placed inertial measure units (IMUs, TDK InvenSense, ICM42607, sampling rate at 100 Hz) on both wrists, feet and shoulders to collect local acceleration, angular rate and Euler angles. We also placed pressure sensors on toe, the first metatarsal bone and heel on both feet to divide gait cycle into four phases, including swing phase, initial contact, midstance and propulsion[53]. For Adam’s forward bending test we collected EMG signals from three pairs of paraspinal muscles beside T3, T8 and L1, upper and lower rectus abdominis muscles. We presented the detailed attachment for EMG and IMU sensors for two motion tasks in Fig. S1 and S2.

All the EMG sensors were placed according to the recommendation of SENIAM**Error! Reference source not found.** and confirmed by palpation. All the sensors were time synchronized by a synchronized trigger and time stamps. The data collection hardware systems were well established in our previous studies[55]-**Error! Reference source not found.**

### EMG features

We processed EMG signals in three workflows. The first one was for extracting spatio-temporal features. We bandpass filtered EMG signals of each channel under the cutoff frequencies of 20 Hz and 300 Hz and then notch filtered EMG signals of each channel at 50 Hz using 4^th^ order Butterworth filter. Then, we calculated (1) the root mean square (RMS) ratios of patients (convex/concave) and healthy controls (left/right) for paraspinal muscles and of all the subjects (left/right) for lower-limb muscles during Adam’s test and each of four phases of level walking; (2) the onset durations from each channel of all the subjects during Adam’s test and each gait cycle of level walking; (3) center of activity from each channel of all the subjects during each gait cycle of level walking.

The second workflow was for extracting spectral features. We bandpass and notch filtered EMG signals of each channel using the same parameter as the first workflow, and performed full-wave rectification, then divided the signals into delta (0-5Hz), alpha(5-15Hz) and beta(15-35Hz) bands. We calculated (1) coherence from each pair of frequency bands across muscles on opposite sides during Adam’s test and each gait cycle of level walking; (2) energy of each frequency band and each EMG channel during Adam’s test and each gait cycle of level walking.

The third workflow was for extracting muscle synergies. We performed the standard muscle synergy extraction method as in our previous study**Error! Reference source not found.** after bandpass, notch filtering and full-wave rectifying EMG signals using the same parameter in the second workflow. Then, we calculated (1) the sparseness of muscle synergy**Error! Reference source not found.**; (2) the similarity of muscle synergies across groups**Error! Reference source not found.**. The detailed methods for calculating the EMG features were presented in SI.

When extracting EMG features for Adam’s forward bending test we discarded the data of the first and last 3 second, and then performed the same procedure except for windowing the signals.

### Kinematic features

We processed IMU signals for all the three groups of subjects during level walking. We bandpass filtered IMU signals of each channel at the cutoff frequencies of 0.1Hz and 15Hz using 4^th^ order Butterworth filter. We calculated the following features: (1) the difference between maximum and minimum of each Euler angle of each IMU channels for each phase of the gait; (2) spatio-temporal gait parameters, including step length, foot progression angle, step width, single and double support phase (%), cadence, the vertical range of motion of center of mass, vertical range of motion of foot for each gait cycle; (3) the asymmetry of wrist from wrist channel for each gait cycle. The parameters were selected according to previous studies**Error! Reference source not found.**. We presented the details of calculating the IMU features in SI.

### Individualized musculoskeletal model

We collected the biplanar radiographic images and kinematics for each patient and developed individualized musculoskeletal model according to the methods in **Error! Reference source not found.**, which has been validated against measured EMG signals. Specifically, the Opensim-based full-body musculoskeletal models for adolescents**Error! Reference source not found.** were adopted. We then captured 10 key landmarks of each vertebra for each patient using a fully automated deep neural network-based approach to identify the position, orientation and height of each vertebra from T1 to L5, then confirmed by manually checking through experienced radiologist and clinicians in scoliosis treatment. We scaled the musculoskeletal model by matching the participants’ height, weight, and body proportions. We adjusted the spinal deformity of musculoskeletal model by the identified position, orientation and height of each vertebra.

We collected the gait data for the participants through simultaneously using optical motion capture system (VICON Motion System Ltd, Oxford, UK, sampling rate at 100Hz) and ground force plates (AMTI, Watertown, USA, sampling rate at 100 Hz). The participants were asked to walk in the instrumented environment for 10 entire gait cycles (defined as gait cycles with ground reaction force of both feet, maker trajectories and covering whole gait). We re-adjusted the skin markers within the musculoskeletal model to accommodate the modeled spinal deformity. The Inverse Kinematics tool in OpenSim (version 6.0) and MATLAB R2024a (MathWorks, Inc., Natick, MA, USA) and Residual Reduction Algorithm were used to optimize the body mass properties of the model by minimizing the mismatch between model-simulated and measured kinematics. The Inverse Dynamics tool and Static Optimization were used to estimate net joint moment and allocate the joint moment to muscle forces. Finally, Joint Reaction Analysis tool was used to estimate internal loads among vertebra.

We estimated the vertebral loading for vertebras from T1 to L5 in terms of loading forces and loading torques between adjacent vertebra in ground coordinate system (defined as X axis: the forward direction of walking, Y axis: vertically upward, Z axis: the right direction of walking). That is, we estimated the loading force in all the three axes and the loading torque surrounding all the three axes of ground coordinate system.

### Statistics

For assessing the distance between two groups of participants A and B (Fig. 2a-c, Fig. 4a and Fig. 5b), we calculated the maximum mean discrepancy (MMD) between two groups and performed bootstrapping to construct distribution of the distance. Specifically, we first stacked the extracted feature vectors (each element corresponded to a specific feature) of IMU and EMG over participants and steps for level walking (or sliding windows for Adam’s test) for each motion task. We then reduced the dimension of features by performing PCA respectively for each motion task and remaining the principal components that can explain over 90% variance of the feature matrix. We randomly sampled 1000 feature vectors (after dimension reduction) from each group of participants with replacement (e.g. for the leftmost bar of Fig. 2b, we sampled 1000 vectors from functional scoliosis patients and another 1000 vectors from healthy controls). Then, we calculated MMD between these two sampled sets of feature vectors. Then, we performed bootstrapping for 1000 times to construct the MMD distribution consisting of 1000 MMD values as the distance between group A and B.

For assessing the regression coefficients, we performed linear regression (Fig. 2, 4 and 5). We stacked the feature vectors in the same setting, performed PCA on feature dimension, and remained the first principal component. Then, we performed regressions using the first principal components for each pair of participants within groups. For example, when we regressed the EMG features of functional scoliosis against those of healthy controls (Fig. 2d), we performed PCA respectively for each group of participants. We regressed the first principal component of one participant within functional scoliosis group against the first principal component of every participant within healthy control group, respectively. We performed the regression for every participant within functional scoliosis group. We thus constructed a distribution of regression coefficients denoting the association between these two groups with 33*18 values (33 functional scoliosis patients and 18 healthy control). Then, we compared across the distributions of regression coefficients between different pairs of groups.

For the regression presented in the left panel of Fig. 5c, we remained the first principal components of vertebral loading and then regressed the screened EMG features (detailed in Machine learning-based analysis) of each subject over all the three groups of participants during level walking against these first principal components.

### Machine learning-based analysis

We leveraged machine learning in two analyses. The first one was to subgrouping functional scoliosis patients into those who presented similar muscle activation patterns with healthy controls and those similar to idiopathic scoliosis patients (Fig. 2e and Fig. 3c). Specifically, we used C-SVM to classify which group (healthy, functional or idiopathic scoliosis) the participant belonged to and performed 5-fold cross-validation on EMG and kinematic features, respectively. We finally used the prediction in the test set of each fold so that decoder made predictions on all the data and then constructed the confusion matrix. We used the first row of the confusion matrix in Fig. 2e and Fig. 3c to divide the functional scoliosis patients in two three subgroups. Then, we compared the associations between each of the subgroups and healthy controls (Fig. 2f, Fig. 4d), as well as the associations between each of the subgroups and idiopathic scoliosis (Fig. 2g and Fig. 4e).

The second analysis was to screen the distinguishable features among healthy controls, functional and idiopathic scoliosis patients (Fig. 4g and Fig. 5c). Specifically, we performed two machine learning algorithms to classify which group (healthy, functional or idiopathic scoliosis) the participant belonged to and then selected the distinguishable features shared across the two algorithms. We used C-SVM and performed the same 5-fold cross-validation paradigm as mentioned above. Then, we shuffled each feature across participants while fixed other features. In this way, we destroyed the correlation between the shuffled feature with other features. We then trained and tested C-SVM in the same manner. We performed the shuffling-training-testing procedure 1000 times for each feature. Then, we assessed the shuffling-induced accuracy degradation by subtracting the mean accuracy after shuffling from the accuracy before shuffling. We selected 10 features with top 10 accuracy degradation. We trained and validated logistic regression in the same 5-fold cross-validation manner where we randomly divided the data into 5 folds again. Then, we used the logistic regression-based decoder with the highest accuracy on validation fold. From this decoder, we selected 10 features with the top 10 weights within the decoder. Finally, we determined the distinguishable features as the shared features across the selected features from the two decoder-based analyses.

### Gene expression analysis

We leveraged two open-sourced datasets on idiopathic scoliosis in this analysis (Fig. 6). The first dataset consisted of mRNA of bilateral paraspinal muscles from 5 idiopathic scoliosis patients whose apex located between T5/6 and T11/12 taken during surgery**Error! Reference source not found.**. The second dataset consisted of circulating micro RNAs from 116 adolescent idiopathic scoliosis patients (mean age 13.3 ± 1.7 years, mean Cobb angle of 24.4° ± 12.4°) and 30 healthy controls[23].

For the first dataset, we also averaged the mRNA data of paraspinal muscles across participants, We then stacked EMG, kinematic features or vertebral loading of level walking over subjects and gait cycles for each of the three groups of participants, respectively. We performed PCA to remain the top 15 principal components according to the variance explained for each of the three groups of the participants recruited in out study, respectively. We calculated the Pearson’s correlation coefficients between the top 15 principal components of the mRNA data averaged across 5 idiopathic scoliosis patients [] and the the top 15 principal components of EMG, kinematic features or vertebral loading of each group of participants recruited in our study. In this way, we evaluated the similarity between the kinematics, muscle activation pattern and vertebral loading pattern of healthy, functional and idiopathic scoliosis participants with the gene expression within paraspinal muscles of idiopathic scoliosis patients.

For the second dataset, we averaged the circulating micro RNA data across subjects within each group (i.e. idiopathic scoliosis and healthy controls). We also stacked the EMG, kinematic features or intervertebral disc loading of level walking in the same way. We performed PCA on the feature dimension and remained the top 15 principal components according to the variance explained for functional scoliosis patients. We calculated the Pearson’s correlation coefficients between the top 15 principal components of circulating microRNA averaged across 30 healthy controls and across 116 idiopathic scoliosis patients and the top 15 principal components of EMG, kinematic features or vertebral loading of each functional scoliosis patient recruited in our study. In this way, we evaluated the similarity between the kinematics, muscle activation pattern and intervertebral disc loading pattern of functional scoliosis with the circulating gene expression of healthy and idiopathic scoliosis participants.

## Data availability

The transcriptomic data used in this study are mRNA of bilateral paraspinal muscles from 5 idiopathic scoliosis patients opensourced by Wang et. al. (https://www.ncbi.nlm.nih.gov/geo/query/acc.cgi?acc=GSE254300) and circulating micro RNAs from 116 adolescent idiopathic scoliosis patients open-sourced by Khatami et. al. (https://www.ncbi.nlm.nih.gov/geo/query/acc.cgi?acc=GSE286204#:~:text=In%20this%20prospective%20crosssectional%20study%2C%20we%20investigated%20the,the%20risk%20of%20developing%20severe%20scoliosis%20in%20AIS).The data used for generating the figures were uploaded to Github upon acceptance. The raw data will be provided upon reasonable request from M.D. Xiaolei Sun.

## Code availability

The code for this study will be made available on Github upon acceptance.

## Acknowledgments

This work was supported by National Natural Science Foundation of China (No. 62306083), Key Research Project funded by Chinese Association of Rehabilitation Medicine (No. KFKT-2024-011) and the China Scholarship Council.

## Author Contributions

C.Y., L.A. and X.S. conceived the project and designed the experiments. All authors contributed to the development of the concepts presented in the paper. Z.H., J.Z., X.S. and B.W. performed the experiments. All authors designed the data analysis approaches. Z.X., B.W. and H.Z. conceived the application of the musculoskeletal model. Z.X., J.Z. B.W. conceived statistical analysis. B.W. performed the data analysis. C.Y. and B.W. wrote the manuscript with contributions from all the rest authors. All authors contributed to the content and writing of the Supplementary Information.

