## Supplemental Information for "The geno-biomechanical similarity between functional and idiopathic scoliosis from kinematic, muscle activation, vertebral loading and the pathological gene expression perspectives"

* The authors share the same contribution;

2. EMG feature extraction
(1) Temporal features

Root mean square (RMS) ratio: For the pre-processed EMG signal x(t) from each channel, we calculated the RMS as follow.

$$\text{RMS}\text{=}\sqrt{\frac{\text{1}}{\text{N}}\sum_{\text{t}\text{=1}}^{\text{N}} \text{x}\left( \text{t} \right)^{\text{2}}}$$

where N denotes the number of sample points within each gait cycle for shuffle walking, within the whole duration except for the first and last 3 seconds for Adam’s forward bending test. Then, the RMS ratio was given by

$$\text{RMS}\text{ }\text{ratio}\text{=}\frac{\text{RM}\text{S}_{\text{left}}}{\text{RM}\text{S}_{\text{rig}\text{h}\text{t}}}$$

where the subscripts, left and right, denote the RMS of the paraspinal muscles on the left and right sides, respectively.

Onset duration: For the pre-processed EMG signal x(t) from each channel, we first got the envelope of EMG $\text{x}_{\text{env}}\left( \text{t} \right)$ by full-wave rectification and then low-pass filter by a 4^th^ order Butterworth filter with the cut-off frequency of 15Hz. Then, we calculated the onset as follows.

$$\text{Onset}\left( \text{t} \right)\text{=}\left\{ \begin{aligned} \text{1, }\text{if}\text{ }\text{x}_{\text{env}}\left( \text{t} \right)\text{>}\text{μ}\text{+3}\text{σ} \\ \text{0, }\text{else} \end{aligned} \right.$$

where $\text{μ}$ denotes the mean of $\text{x}_{\text{env}}\left( \text{t} \right)$ within each gait cycle for shuffle walking and within the whole duration except for the first and last 3 seconds for Adam’s forward bending test, $\text{σ}$ denotes the standard deviation of $\text{x}_{\text{env}}\left( \text{t} \right)$ within each gait cycle for shuffle walking and within the whole duration except for the first and last 3 seconds for Adam’s forward bending test. The onset duration is then given by

$$\text{D}\text{=}\frac{\text{1}}{\text{N}}\sum_{\text{t}\text{=1}}^{\text{N}} \text{Onset}\text{(}\text{t}\text{)}$$

where N denotes the number of sample points within each gait cycle for shuffle walking, within the whole duration except for the first and last 3 seconds for Adam’s forward bending test.

Center of Activity (CoA): This feature was only calculated from each channel of all the subjects during each gait cycle of level walking. We first normalized gait cycle into radians, i.e. each gait cycle corresponded to the duration [0, 1]. CoA was then calculated as follows.

$$\text{θ}\text{=arctan(}\frac{\sum_{\text{t}\text{=1}}^{\text{N}} \sin\left( \frac{\text{2}\text{πt}}{\text{N}} \right)\text{x}_{\text{env}}\text{(}\text{t}\text{)}}{\sum_{\text{t}\text{=1}}^{\text{N}} \cos\left( \frac{\text{2}\text{πt}}{\text{N}} \right)\text{x}_{\text{env}}\text{(}\text{t}\text{)}}\text{)}$$

where t denotes the timing within each gait cycle falling into the duration [0,1]. In this way, CoA denotes the main activation phase within a gait cycle for each muscle.

(2) Spectral features

We first divided EMG envelope,${\text{ }\text{x}}_{\text{env}}\text{(}\text{t}\text{)}$, into four frequency bands, i.e. δ (0–5 Hz)，α (5–15 Hz)，β (15–35 Hz).

Energy of each frequency band: We calculated the frequency energy by

$$\text{P}_{\text{b}}\text{=}\frac{\text{1}}{\text{N}}\sum_{\text{t=1}}^{\text{N}} \text{x}_{\text{b}}\left( \text{t} \right)^{\text{2}}$$

where $\text{x}_{\text{b}}\text{(t)}$ denotes the time series within each frequency band, b∈{δ,α,β}, N denotes the number of sample points within each gait cycle for shuffle walking, within the whole duration except for the first and last 3 seconds for Adam’s forward bending test.

Coherence: We calculated coherence from each pair of frequency bands across muscles on opposite sides during Adam's forward bending test (the whole duration after discarding the first and last 3 seconds) and each gait cycle of level walking, given by

$$\text{C}_{\text{ij}}^{\text{(}\text{b}_{\text{1}}\text{,}\text{b}_{\text{2}}\text{)}}\text{=}\text{corr}\text{(}\text{x}_{\text{i}}^{\text{b}_{\text{1}}}\text{,}\text{x}_{\text{j}}^{\text{b}_{\text{2}}}\text{)}$$

where i and j denote the muscles on opposite sides, $\text{b}_{\text{1}}$ and $\text{b}_{\text{2}}$ denote frequency bands, $\text{b}_{\text{1}}$, $\text{b}_{\text{2}}$∈{δ,α,β}.

(3) Muscle synergy

We extracted muscle synergy from EMG envelope,${\text{ }\text{x}}_{\text{env}}\text{(}\text{t}\text{)}$, through non-negative matrix decomposition (NMF)[1].

$$\text{M}\text{=}\text{W}\text{∙}\text{P}\text{+}\text{e}$$

where $\text{M}\text{∈}\text{R}^{\text{m}\text{×}\text{T}}$ denotes the matrix of EMG envelope with m muscles and T sample points, $\text{W}\text{∈}\text{R}^{\text{m}\text{×}\text{N}}$ denotes the muscle synergy matrix with N muscle synergies, $\text{P}\text{∈}\text{R}^{\text{N}\text{×}\text{T}}$ denotes the activation of muscle synergies, $\text{e}$ denotes the residual error matrix. We determined the optimal N as follows. We performed parameter sweep for N from N=2 to N=8. For each N, we calculated the variance accounted for (VAF) as

$$\text{VAF}\text{=1−}\frac{\sum\left( \text{M}\text{−}\hat{\text{M}} \right)^{\text{2}}}{\sum\text{M}^{\text{2}}}$$

where $\hat{\text{M}}$ denotes the estimated muscle activation given by $\hat{\text{M}}\text{=}\text{W}\text{∙}\text{P}$, $\text{Σ}$ denotes summing over every element of a matrix. We determined the optimal N by the highest VAF (at least VAF≥80%).

The similarity of muscle synergies: We regressed across muscle synergy matrices[2], given by

$$\text{W}_{\text{i}}^{\text{a}}\text{=}\sum_{\text{j}\text{=1}}^{\text{N}} \text{m}_{\text{ij}}\text{∙}\text{W}_{\text{j}}^{\text{b}}$$

where $\text{W}_{\text{i}}^{\text{a}}$ denotes the *i*th muscle synergy (i.e. the *i*th column vector of the muscle synergy matrix W) of the *a*th subject, $\text{W}_{\text{j}}^{\text{b}}$ denotes the the *j*th muscle synergy (i.e. the *j*th column vector of the muscle synergy matrix W) of the *b*th subject, $\text{m}_{\text{ij}}$ denotes the regression coefficient calculated by non-negative least squares fit. We averaged across i and j for each pair of subjects and then performed statistics.

The sparsness of muscle synergies: This feature was to quantify the number of active muscles during motion tasks. We calculated the spareness $\text{φ}_{\text{i}}$ by

$$\text{φ}_{\text{i}}\text{=}\frac{\sqrt{\text{N}}\text{−}\frac{\sum_{\text{k=1}}^{\text{N}} \left| \text{W}_{\text{i}}^{\text{k}} \right|}{\sqrt{\sum_{\text{k=1}}^{\text{N}} {\text{(}\text{W}_{\text{i}}^{\text{k}}\text{)}}^{\text{2}}}}}{\sqrt{\text{N}}\text{−1}}$$

where $\text{W}_{\text{i}}^{\text{k}}$ denotes the kth element of the ith muscle synergy (i.e. the kth muscle component of this muscle synergy), N denotes the number of muscle synergies, $\text{φ}_{\text{i}}$ denotes the sparseness of the ith muscle synergy. We then averaged across muscle synergies for each subject and then performed statistics.

3. Kinematic features

We selected kinematic features according to[3]. All the features were calculated using the IMU signals after preprocessing (see “Methods-Kinematic features”).

(1) Temporal features

Cadence: We counted steps using the foot pressure signals and then calculated cadence by

$$\text{Cadence=}\frac{\text{N}_{\text{steps}}}{\text{T}_{\text{total}}}$$

where $\text{N}_{\text{steps}}$ denotes the steps performed for this subject, $\text{T}_{\text{total}}$ denotes the time (second) used for performing such steps.

Double support ratio: We got the double support ratio for each gait cycle using foot pressure signals by

$$\text{Double support ratio=}\frac{\text{T}_{\text{support}}}{\text{T}_{\text{cycle}}}$$

where $\text{T}_{\text{support}}$ denotes the time of double support phase within this gait cycle, $\text{T}_{\text{cycle}}$ denotes the time for the whole gait cycle. We got one value for this feature for each gait cycle.

Swing ratio: We got the swing ratio for each gait cycle using foot pressure signals by

$$\text{Swing Ratio=}\frac{\text{T}_{\text{swing}}}{\text{T}_{\text{cycle}}}$$

where $\text{T}_{\text{swing}}$ denotes the time of swing phase within each gait cycle. We got one value for this feature for each gait cycle.

(2) Spatial features

Asymmetry of wrist: We used this feature to represent the asymmetry of shoulder during walking between left and right sides according the wrist IMUs. We first calculated the intensity of hand swing using the IMU signals in the transversal plane[4], given by

$$\text{I}_{\text{s}}\left[ \text{n} \right]\text{=}\text{std}\left( \sqrt{\text{S}_{\text{x}}\left[ \text{n} \right]^{\text{2}}\text{+}\text{S}_{\text{z}}\left[ \text{n} \right]^{\text{2}}} \right)$$

where $\text{S}_{\text{x}}\left[ \text{n} \right]$ and $\text{S}_{\text{z}}\left[ \text{n} \right]$ denote the X and Z axes of preprocessed IMU signals, respectively, $\text{std}\text{( )}$ denotes the standard deviation. We then calculated the asymmetry of wrist by

$$\text{Feature}_{\text{asym}}\text{=}\frac{\text{Intensity}_{\text{left}}}{\text{Intensity}_{\text{rig}\text{h}\text{t}}}$$

Displacement of waist and feet in Z axis: We calculated these features within each gait cycle. We segmented the gait cycles using foot pressure sensors and integrated the acceleration in the Z axis since heel strike within each gait cycle. We performed the dual integration by

$$\text{v}_{\text{z}}\text{(}\text{t}\text{)=}\frac{\text{1}}{\text{f}_{\text{s}}}\sum_{\text{τ}\text{=1}}^{\text{t}} \text{ }\text{a}_{\text{z}}\text{(}\text{τ}\text{)}$$

where $\text{a}_{\text{z}}\text{(}\text{τ}\text{)}$ denotes the sample point of the Z axis of IMUs mounted on feet and waist, $\text{f}_{\text{s}}$ denotes the sampling frequency. We then calculated the displacement by

$$\text{d}_{\text{z}}\text{(}\text{t}\text{)\&=}\frac{\text{1}}{\text{f}_{\text{s}}}\sum_{\text{τ}\text{=1}}^{\text{t}} \text{ }\text{v}_{\text{z}}\text{(}\text{τ}\text{)}$$

We then calculated these features as

$$\text{Feature}_{\text{displacement}}\text{=}\text{d}_{\text{z}}\text{(}\text{T}\text{)}$$

where T denotes the end of a gait cycle.

Foot progression angle during foot flat: We calculated the foot progression angle according to the method presented in []. Specifically, we first calculated the axis of plantar flexion by the angular velocity averaged between heel strike and foot flat and then normalized to unit magnitude, denoted by $\text{k}^{\text{s}}\text{.}$

$$\text{k}^{\text{s}}\text{=}\left\{ \begin{aligned} \text{k}^{\text{s}}\text{, }\text{if}\text{ }\text{k}^{\text{s}}\text{(}\text{x}\text{) > 0 }\text{for}\text{ }\text{left}\text{ }\text{foot}\text{ } \\ \begin{matrix} \text{k}^{\text{s}}\text{, }\text{if}\text{ }\text{k}^{\text{s}}\text{(}\text{x}\text{) < 0 }\text{for}\text{ }\text{rig}\text{h}\text{t}\text{ }\text{foot} \\ \text{−}\text{k}^{\text{s}}\text{, }\text{ot}\text{h}\text{erwise} \end{matrix} \end{aligned} \right.$$

where $\text{k}^{\text{s}}\text{(}\text{x}\text{)}$ denotes the X axis of $\text{k}^{\text{s}}$. We then calculated the foot progression direction as

$$\text{f}^{\text{S}}\text{=}\text{k}^{\text{s}}\text{×}\text{g}^{\text{S}}$$

where S denotes the coordinate frame of foot-mounted IMU, $\text{g}^{\text{S}}$ denotes the vertical direction calculated by averaging the acceleration across foot flat and normalizing to unit magnitude. We then transformed the foot progression direction into inertial coordinate frame using the Euler angles calculated by the Kalman filter embeded within the IMU sensors, denoted by $\text{f}^{\text{E}}$.

We then calculated the displacement of the foot in the horizontal plane by integrating the acceleration within inertial coordinate frame in the horizontal plane from foot flat to heel strike, given by

$$\text{r}_{\text{xy}}\text{=}\int\int_{\text{foot}\text{ }\text{flat}}^{\text{h}\text{eel}\text{ }\text{strike}} \text{a}_{\text{xy}}^{\text{E}}\text{(}\text{t}\text{)}\text{dtdt}$$

where $\text{a}_{\text{xy}}^{\text{E}}\text{(}\text{t}\text{)}$ denotes the readings of X and Y axes of the accelerometer in the inertial coordinate frame at sampling point t. We then calculated the foot progression angle during foot flat as

$$\text{θ}_{\text{progress}}\text{=arccos}\left( \frac{\text{f}_{\text{xy}}^{\text{E}}\text{⋅}\text{r}_{\text{xy}}}{\text{∥}\text{f}_{\text{xy}}^{\text{E}}\text{∥⋅∥}\text{r}_{\text{xy}}\text{∥}} \right)\text{×}\frac{\text{180}}{\text{π}}$$

Step length: We projected the displacement of the foot, $\text{r}_{\text{xy}}$, into the direction of foot progression in the horizontal plane, given by

$$\text{L}_{\text{step}}\text{=}\text{r}_{\text{xy}}\text{⋅}\text{f}_{\text{xy}}^{\text{E}}$$

Step width: We projected the displacement of the foot, $\text{r}_{\text{xy}}$**,** in the horizontal plane into the direction perpendicular to the foot progression and vertical directions, given by

$$\begin{aligned} \text{l}\text{=}\text{f}^{\text{E}}\text{×[0,0,−1}\text{]}^{\text{T}} \\ \text{W}_{\text{step}}\text{=}\text{r}_{\text{xy}}\text{⋅}\text{l}_{\text{xy}} \end{aligned}$$

where $\text{l}_{\text{xy}}$ denotes the X and Y axes of the vector $\text{l}$.

Range of motion: We calculated the difference between the maximum and minimum of Euler angles of each IMU during each gait cycle by

$$\text{Feature}_{\text{euler}}\text{=[}\text{max}\text{(}\text{roll}\text{)−}\text{min}\text{(}\text{roll}\text{),max(}\text{pitc}\text{h)−}\text{min}\text{(}\text{pitc}\text{h),max(}\text{yaw}\text{)−}\text{min}\text{(}\text{yaw}\text{)]}$$

*Relate to the photo of subjects. Please contact the authors for details.*

**Fig. S1** The schematic diagram of EMG and IMU placement. The EMG sensors were placed on paraspinal and lower-limb muscles for shuttle walking, and the IMU sensors were placed on shoulders, wrists and feet (**a** and **c**). For Adam’s forward bending test, we placed EMG sensors on paraspinal, abdominal, gluteal and thigh muscles (**b** and **d**).

*Relate to the photo of subjects. Please contact the authors for details.***Fig. S2** The schematic diagram of IMU placement and the orientations of the sensor coordinate frames.

*Relate to the photo of subjects. Please contact the authors for details.*

**Fig. S3** The schematic diagram of placing reflective markers for the optical motion capture system and ground force plates.

**Tab. S1 The demographic information of recruited functional scoliosis patients**

| Number | Gender | Cobb Angle(°) | Curve Pattern |
| --- | --- | --- | --- |
| 1 | Female | 11 | Lumbar, left |
| 2 | Male | 11 | Lumbar, left |
| 3 | Female | 11 | Lumbar, left |
| 4 | Female | 10 | Lumbar, left |
| 5 | Female | 10 | Thoracic, right |
| 6 | Male | 12 | Thoracic, left |
| 7 | Male | 11 | Lumbar,right |
| 8 | Female | 11 | Thoracic, left |
| 9 | Male | 12 | Lumbar, right |
| 10 | Male | Thoracic: 13 Lumbar: 7 | Thoracic, left  Lumbar, right |
| 11 | Female | Thoracic: 15  Lumbar: 8 | Thoracic, right  Lumbar, left |
| 12 | Male | 10 | Thoracic, left |
| 13 | Male | Thoracic: 8 Lumbar: 8 | Lumbar, left |
| 14 | Male | 11 | Lumbar, left |
| 15 | Female | 13 | Lumbar, left |
| 16 | Male | 10 | Lumbar, left |
| 17 | Male | 11 | Lumbar, left |
| 18 | Female | 11 | Lumbar, left |
| 19 | Male | 15 | Thoracic-lumbar (kyphosis) |
| 20 | Female | Thoracic: 13  Lumbar: 12 | Thoracic,right lumbar, left |

**Tab. S2 The demographic information of recruited idiopathic scoliosis**

| Number | Gender | Cobb Angle(°) | Curve Pattern |
| --- | --- | --- | --- |
| 1 | Female | 20 | Lumbar, right |
| 2 | Female | Thoracic: 26  Lumbar: 35 | Thoracic, left  Lumbar, right |
| 3 | Male | Thoracic: 18  Lumbar: 20 | Thoracic, left  Lumbar, right |
| 4 | Female | Thoracic: 26  Lumbar: 18 | Thoracic, right  Lumbar, left |
| 5 | Female | Thoracic: 12  Lumbar: 28 | Thoracic, right  Lumbar, left |
| 6 | Female | Thoracic: 13  Lumbar: 16 | Thoracic, left  Lumbar, right |
| 7 | Female | 22 | Lumbar, left |
| 8 | Female | 23 | Lumbar, left |
| 9 | Male | Thoracic: 24  Lumbar: 22 | Thoracic, left  Lumbar, right |
| 10 | Female | 19 | Thoracic, right |
| 11 | Male | 20 | Thoracic, left |
| 12 | Male | Thoracic: 23  Lumbar:18 | Thoracic, left  Lumbar, right |
| 13 | Female | Thoracic: 15  Lumbar: 16 | Thoracic,right  Lumbar, left |
| 14 | Female | Thoracic: 25  Lumbar: 23 | Thoracic, left  Lumbar, right |
| 15 | Female | 17 | Lumbar, left |
| 16 | Female | Thoracic: 18  Lumbar: 21 | Thoracic, left  Lumbar, right |
| 17 | Male | Thoracic: 7  Lumbar: 21 | Thoracic, left  Lumbar, right |
| 18 | Female | 17 | Lumbar, left |
| 19 | Female | 19 | Lumbar, left |
| 20 | Female | Thoracic: 21  Lumbar: 24 | Thoracic, left  Lumbar, right |
| 21 | Female | Thoracic: 30  Lumbar: 10 | Thoracic, left  Lumbar, right |

**Tab. S3 The demographic information of recruited healthy subjects**

| Number | Gender |
| --- | --- |
| 1 | Male |
| 2 | Male |
| 3 | Male |
| 4 | Male |
| 5 | Female |
| 6 | Female |
| 7 | Male |
| 8 | Female |
| 9 | Male |
| 10 | Female |
| 11 | Male |
| 12 | Female |
| 13 | Female |
| 14 | Male |
| 15 | Male |
| 16 | Female |
| 17 | Male |
| 18 | Male |
